# Longitudinal changes in iron homeostasis in human experimental and clinical malaria

**DOI:** 10.1101/2023.12.19.23300265

**Authors:** Stephen D. Woolley, Matthew J. Grigg, Louise Marquart, Jeremy Gower, Kim Piera, Arya Sheela Nair, Fiona M. Amante, Giri S. Rajahram, Timothy William, David M. Frazer, Stephan Chalon, James S. McCarthy, Nicholas M. Anstey, Bridget E. Barber

## Abstract

**Background:** The interaction between iron deficiency and malaria is incompletely understood. We evaluated longitudinal changes in iron homeostasis in volunteers enrolled in malaria volunteer infection studies (VIS) and in Malaysian patients with falciparum and vivax malaria.

**Methods:** We retrieved samples and associated data from 55 participants enrolled in malaria VIS, and 171 malaria patients and 30 healthy controls enrolled in clinical studies in Malaysia. Ferritin, hepcidin, erythropoietin, and soluble transferrin receptor (sTfR) were measured by ELISA.

**Results:** In the VIS, participants’ parasitaemia was correlated with baseline mean corpuscular volume (MCV), but not iron status (ferritin, hepcidin or sTfR). Ferritin, hepcidin and sTfR all increased during the VIS. Ferritin and hepcidin normalised by day 28, while sTfR remained elevated. In VIS participants, baseline iron status (ferritin) was associated with post-treatment increases in liver transaminase levels. In Malaysian malaria patients, hepcidin and ferritin were elevated on admission compared to healthy controls, while sTfR increased following admission. Hepcidin normalised by day 28; however, ferritin and sTfR both remained elevated 4 weeks following admission.

**Conclusion:** Our findings demonstrate that parasitaemia is associated with an individual’s MCV rather than iron status. The persistent elevation in sTfR 4 weeks post-infection in both malaria VIS and clinical malaria may reflect a causal link between malaria and iron deficiency.

## Introduction

Malaria and iron deficiency are major public health problems worldwide, with both frequently co-existing in vulnerable populations. Despite the significant global prevalence of both conditions, the interaction between iron deficiency and malaria is incompletely understood. Iron deficiency has been shown to protect against clinical malaria in African children^1^ and pregnant women,^2^ and in African children protects against severe malaria and death.^3^ Furthermore, iron supplementation has been associated with subsequent malaria infection and mortality.^4^ These clinical studies are supported by *in vitro* data demonstrating that *P. falciparum* infects iron-deficient erythrocytes less efficiently, and that this effect is reversed by iron supplementation.^5^ However, the interaction between an individual’s iron status and parasite growth *in vivo* has not been defined.

The impact of malaria infection on iron deficiency has also not been fully characterised. Recently, a causal link between malaria and iron deficiency was suggested by a Mendelian randomization analysis that demonstrated that the sickle cell trait (protective against malaria) was associated with a reduced prevalence of iron deficiency in African children.^6^ Whether this causal link occurs in other populations, and the underlying mechanisms, has not been determined.

Characterising the complex interaction between malaria and iron deficiency is further hindered by the difficulties in interpreting the various markers of iron status. Iron metabolism is controlled by the hepatocyte produced hormone hepcidin, which binds to and degrades the iron transmembrane exporter ferroportin.^7–9^ Hepcidin therefore reduces iron availability by preventing iron export from the reticuloendothelial system, enterocytes and hepatocytes, subsequently inducing a hypoferraemia.^7–9^ Hepcidin is upregulated by interleukin-6 (IL-6) via the JAK/STAT3 pathway,^10^ and has been shown to be increased in African children with falciparum malaria,^11,12^ and in adults with vivax malaria.^13^ Ferritin, the key intracellular iron storage protein but also an acute phase protein, is also elevated in malaria.^14,15^ Soluble transferrin receptor (sTfR) has been proposed as a more reliable indicator of iron status as it is not affected by inflammation.^8,16^ However, synthesis of sTfR is also increased by erythropoiesis and haemolysis. Thus, while elevated sTfR levels have been observed in children with malaria,^14,17,18^ whether this reflects a functional iron deficiency, erythropoiesis or haemolysis, remains uncertain.

To better understand the complex interaction between *Plasmodium* infection and iron homeostasis, we evaluated the longitudinal changes in hepcidin, ferritin and sTfR in volunteers experimentally infected with blood-stage *P. falciparum*, and in Malaysian patients (mostly adults) with falciparum and vivax malaria. We sought to determine 1) whether there is an association between an individual’s iron status and parasite growth, and 2) the impact of experimental and clinical malaria infection on iron homeostasis in adults. Finally, given previous observations of abnormal liver function tests following antimalarial treatment in both malaria volunteer infection studies (VIS)^19,20^ and clinical malaria,^21^ and the importance of this finding for antimalarial drug development, we investigated whether this adverse finding in malaria VIS was associated with participants’ baseline iron status.

## Methods

### Malaria volunteer infection studies (VIS)

Data were retrieved from the records of 55 participants enrolled in 7 induced blood stage malaria (IBSM) VIS conducted in Brisbane, Australia, between 2015 and 2021.^22–26^ Study participants were malaria-naïve adults aged 18-55 years with no significant medical co-morbidities or concurrent illness. Participants were inoculated with erythrocytes parasitized with either the *P. falciparum* 3D7 strain (n=45), or the artemisinin-resistant *P. falciparum* K13 strain (n=10). Parasitaemia was closely monitored at specific time-points using quantitative PCR (qPCR) targeting the species specific 18S ribosomal ribonucleic (rRNA) gene. The studies evaluated six different antimalarial drugs (including registered antimalarials and new chemical entities in development), with the day of administration being day 9 for the K13 *P. falciparum* strain and day 8 for the 3D7 strain. For many of the studies, sub-curative doses of the antimalarial drugs were intentionally administered, resulting in parasite recrudescence to allow for the calculation of drug minimum inhibitory concentrations. All participants dosed with a new chemical entity were treated with a registered antimalarial drug such as artemether-lumefantrine or atovaquone-proguanil, at the time of recrudescence or at the end of study.

### Parasite data and laboratory measurements

Parasitaemia parameters evaluated in the VIS participants included the log10 transformed peak pre-treatment parasitaemia (parasites/mL), the total parasite burden (area under the curve of the non-transformed 18S qPCR data from day 4 until time of treatment), and the parasite multiplication rate (PMR), which have all been previously reported.^26^ All VIS participants had blood collected for standard haematology (including reticulocyte count) and biochemistry prior to inoculation (day 0), prior to antimalarial treatment (day 8 or 9), on day 15 (+/-2), and at the end of study (EOS, day 30 +/-3). The reticulocyte difference was defined as the difference between the reticulocyte count at EOS and the reticulocyte count prior to inoculation.

Stored frozen aliquots of serum from the studies were tested using commercial ELISA assays for erythropoietin (EPO; BioLegend), hepcidin (Intrinsic Lifesciences) and sTfR (BioVendor), with all ELISAs performed as per manufacturer’s instructions. If CRP and ferritin levels had not been measured at the appropriate timepoints in the clinical trial, they were tested by immunoturbidimetric (CRP) assay and a 2-step chemiluminescent microparticle (ferritin) immunoassay via an external NATA-accredited clinical laboratory using stored frozen serum. As ferritin is an acute phase protein, its level was adjusted for CRP. The adjustment was based on the Biomarkers Reflecting Inflammation and Nutritional Determinants of Anaemia (BRINDA) project,^27,28^ with ferritin levels remaining unchanged if CRP was ≤5 mg/ml, or multiplied by 0.67 if CRP was >5 mg/mL. Participants were defined as iron deficient if they had an adjusted ferritin below 15 ng/mL.

### Malaysian study sites and study procedures

Patients hospitalised with malaria were enrolled as part of observational studies and/or clinical trials conducted at 4 study sites in Sabah, Malaysia, including a tertiary hospital (Queen Elizabeth Hospital; QEH), and 3 district hospitals (Kudat, Kota Marudu and Pitas District Hospitals) between 2010-2016.^29–33^ For the current analysis only patients with PCR-confirmed falciparum or vivax malaria were included. Patients admitted to the QEH were aged >12 years, were within 18 hours of commencing antimalarial treatment, not pregnant and had no major medical co-morbidities. Inclusion criteria at the district hospitals were similar, except patients were all enrolled pre-treatment, and all ages were included. Treatments given were based on local hospital guidelines at the time, including artemisinin-combination (ACT) for uncomplicated *P. falciparum,* ACT or chloroquine plus primaquine for uncomplicated vivax malaria, and intravenous artesunate for severe malaria. Patients enrolled in the clinical trials were randomised to receive either ACT or chloroquine, according to the study protocols.^31–33^ Healthy controls were visitors or relatives of the study participants admitted to QEH, with no history of fever within 48 hours and blood film negative for malaria.

Venous blood was collected in lithium heparin blood tubes and centrifuged, with plasma stored at -70°C within 30 minutes. For malaria patients admitted to the district hospitals, haematology, biochemistry, and parasite parameters were obtained on enrolment (day 0), and 7 and 28 days following admission. Those admitted to QEH only had samples taken at day 0 and day 28. Stored aliquots of plasma, collected at the above timepoints were tested as described above for hepcidin, ferritin and sTfR levels. The ferritin was not adjusted for CRP as CRP was not available for these patients.

### Ethics Statement

All studies were conducted in accordance with the Declaration of Helsinki and the International Committee of Harmonisation of Good Clinical Practice guidelines. All participants provided informed written consent. For the malaria volunteer infection studies, ethics approval was obtained from the Human Research Ethics Committee of the QIMR Berghofer Medical Research Institute. For the clinical studies in Malaysia, ethics approval was obtained from the Malaysian Ministry of Health and the Menzies School of Health Research, Darwin, Australia. Informed written consent was provided by all participating adults and parents/guardians of those under 18 years of age.

### Statistical analysis

Data were analysed using Stata V17.0 and GraphPad Prism V9.1.0. Normally distributed continuous variables were summarised using the mean and standard deviation (SD), and differences between groups were compared using the Student’s t-test. Non-normally distributed continuous variables were summarised using the median and interquartile range (IQR), and differences between groups compared using the Wilcoxon rank-sum test. Spearman’s correlation was used to investigate the correlations between variables. Predictors of key outcomes of interest in the VIS including total parasite burden (as a measure of parasite growth), peak hepcidin, EOS sTfR and day 15 alanine transferase (ALT) were assessed in separate multivariable linear regression models of biologically plausible factors (log10 transformed due to being non-normally distributed) using backward stepwise elimination of non-significant (p>0.20) factors until all remaining factors were significant (p<0.05). Longitudinal measurements in the malaria VIS were analysed using a repeated measures ANOVA, with non-normally distributed variables log_10_ transformed; the Geisser and Greenhouse correction with a cut-off of p<0.05 was used to determine sphericity. For the longitudinal measurements in the Malaysia studies, a mixed effects model was used to account for missing observations at certain timepoints. All p-values for comparisons between timepoints were obtained from the repeated measures ANOVA (for the malaria VIS) or the mixed effects model (for the Malaysian studies).

## Results

### Malaria volunteer infection studies Demographics and parasite parameters

A total of 55 participants were enrolled in the malaria VIS. The median age of participants was 26 years (IQR 22-35), and 19 (35%) were female (**Table 1**). Compared to males, at baseline females had lower ferritin (median 46 [IQR 22-75] vs 101 [IQR 56-171] ng/mL, p=0.001), lower baseline sTfR (median 1.39 (IQR 1.12 – 1.90) vs 2.01 (IQR 1.65 – 2.43) *μ*g/mL, p=0.002), and higher mean corpuscular volume (MCV; median 89 (IQR 85 – 92) vs 86 (IQR 84 – 88) *μ*m^3^, p=0.011). Only one male participant, and no female, was iron deficient at baseline, defined as a ferritin <15 ng/ml. Participants had a median pre-treatment peak parasitaemia of 16,973 parasites/mL [IQR 4,753-57,602], and a median total pre-treatment parasite burden of 70,920 parasites/mL [IQR 26,017-255,370], with no differences between males and females. There was no correlation between either baseline ferritin or sTfR and peak pre-treatment parasitaemia, total pre-treatment parasite burden, or parasite multiplication rate (**Table 2, Supplementary Table 1**). However, peak pre-treatment parasitaemia and total pre-treatment parasite burden were both associated with baseline MCV (r=0.35, p=0.009; and r=0.39, p=0.004, respectively). The association between baseline MCV and total pre-treatment parasite burden remained significant on multivariable analysis (p=0.005, **Table 2**).

**Table 1.** Clinical and laboratory characteristics of volunteers experimentally infected with *P. falciparum*. Variables are median (interquartile range) unless otherwise stated. Pf: *P. falciparum*; PP_Pre_: peak parasitaemia pre-treatment; SD: standard deviation; TPB_Pre_: total parasite burden pre-treatment; sTFR: soluble transferrin receptor; EPO: erythropoietin; MCV: mean corpuscular volume. *The ferritin was adjusted for inflammation as per the BRINDA project.^27,28^

|  | All (n=55) | Male (n=36) | Female (n=19) | P-value |
| --- | --- | --- | --- | --- |
| Age, years | 26<br>(22-36) | 25<br>(22-32) | 31<br>(22-40) | 0.17 |
| PP<sub>Pre</sub>, parasites/mL | 16,973<br>(4,753-57,602) | 11,674<br>(3,646-31,620) | 32,898<br>(9,062-74,773) | 0.054 |
| TPB<sub>Pre</sub>, parasites | 70,920<br>(26,017-255,370) | 51,978<br>(24,997-247,779) | 138,073<br>(29,987-255,370) | 0.29 |
| Baseline haemoglobin, g/L | 145<br>(136-152) | 149<br>(145-154) | 133<br>(127-138) | **<0.001** |
| End of study haemoglobin, g/dL | 136<br>(127-143) | 141<br>(135-145) | 121<br>(116-131) | **<0.001** |
| Baseline ferritin*, ng/mL | 73 (66-82) | 101 (56-171) | 46 (22-75) | **0.001** |
| End of study ferritin*, ng/mL | 78 (61-76) | 94 (63-179) | 42 (14-63) | **<0.001** |
| Baseline sTfR, µg/mL, (mean, SD) | 1.87 (0.66) | 2.01 (0.63) | 1.50 (0.55) | **0.002** |
| End of study sTfR, µg/mL, | 2.27 (2.17-2.36) | 2.40 (2.10-2.86) | 1.87 (1.63-2.52) | **0.010** |
| Baseline EPO, mIU/mL | 5.71 (5.12-6.36) | 5.75 (3.91-8.94) | 4.44 (3.17-11.16) | 0.60 |
| End of study EPO, mIU/mL | 7.69 (6.90-8.57) | 6.69 (4.34-12.61) | 6.86 (5.32-9.62) | 0.99 |
| Baseline hepcidin, ng/mL | 25 (22-29) | 28 (15-44) | 23 (12-42) | 0.50 |
| Day 8 hepcidin, ng/mL | 30 (26-34) | 33 (19-53) | 32 (10-67) | 0.98 |
| Baseline MCV, fL | 86 (85-89) | 86 (84-88) | 89 (85-92) | **0.010** |
| End of study MCV, fL (mean, SD) | 87 (4.2) | 86 (3.7) | 88 (4.6) | **0.013** |

**Table 2.**
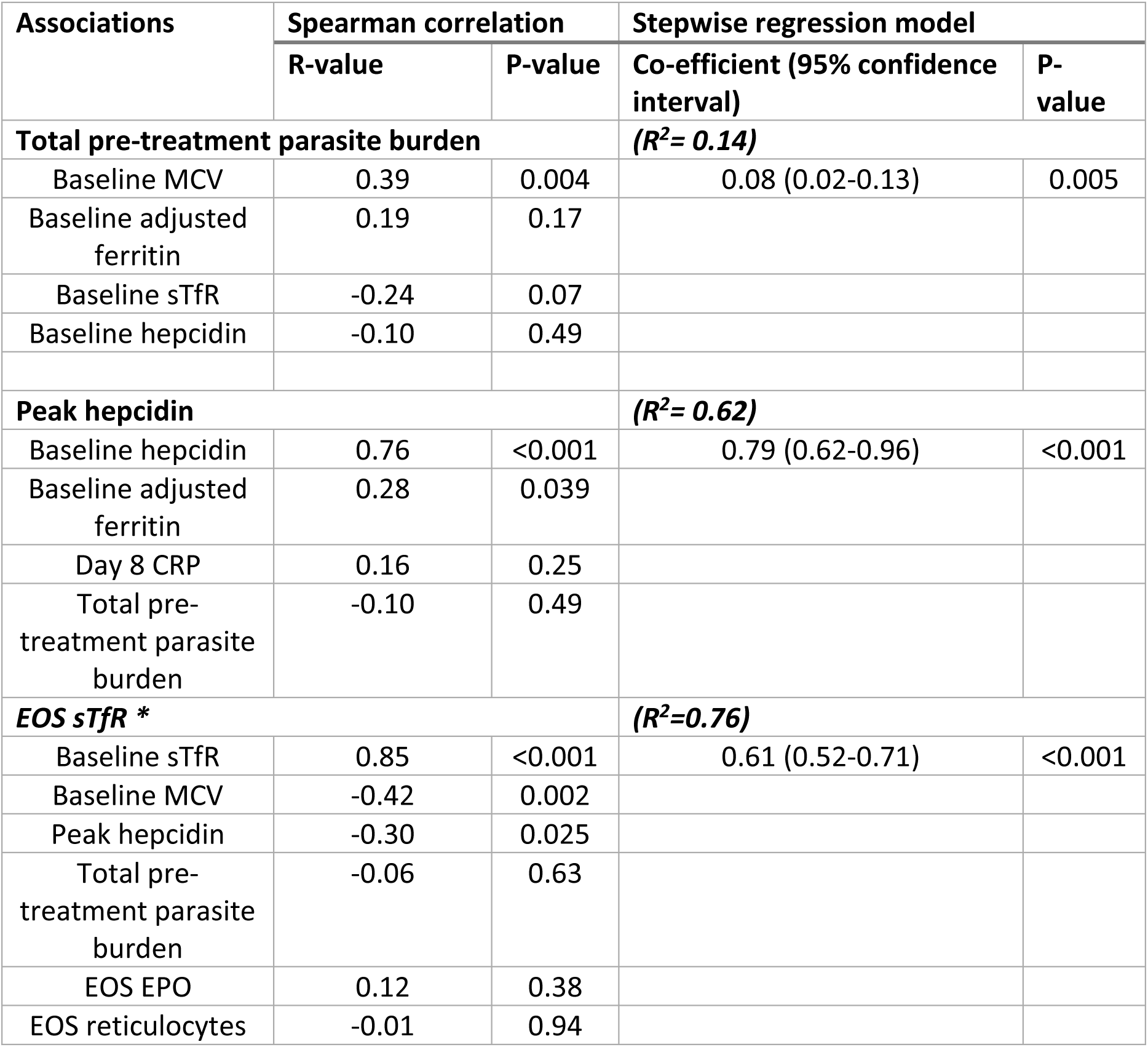
Associations between markers of iron metabolism and parasite parameters in volunteers experimentally infected with *P. falciparum*. Multivariable linear regression analysis of log10 transformed biological plausible factors using backward stepwise elimination. Non-significance for inclusion in the regression model was deemed p>0.20, and variables eliminated until all variables were significant at p<0.05. R^2^: coefficient of determination of the model; EOS: end of study; EPO: erythropoietin; MCV: mean corpuscular volume; sTfR: soluble transferrin receptor. *n=39

### Longitudinal changes in iron markers

Hepcidin rose following inoculation, peaking at day 8, before falling to baseline levels by day 15 (**Figure 1**). Ferritin levels remained unchanged between baseline (median 73 ng/mL [IQR 66-82]) and day of treatment, but then increased following treatment to a median 101 ng/mL (IQR 91-114) on day 15 (p<0.001 for day 8 vs day 15), before returning to baseline levels by day 30. Median sTfR also remained unchanged from inoculation to the day of treatment but increased following treatment from 1.70 *μ*g/mL (IQR 1.63-1.78) to 1.97 *μ*g/mL (IQR 1.88-2.05) at day 15, increasing further to 2.27 *μ*g/mL (IQR 2.17-2.36) by day 30 (p<0.001 for day 8 vs day 30). Like sTfR, median EPO remained stable between baseline and day 8 but increased significantly between day 8 and day 15, and then increased further between day 15 and day 30 (p<0.001 for day 8 vs day 15). Median CRP increased slightly from 1.5 mg/L (IQR 0.3-2.7 mg/L) at baseline to 3.6 mg/L (IQR 2.4-4.8 mg/L; p=0.22) on day 8, increasing further to 9.3 mg/L (IQR 8.1-10.5 mg/L) on day 15 (p<0.001 for day 8 vs day 15), before returning to baseline by EOS. By day 30, 6 participants (11%) met the criteria for iron deficiency, including the single participant who had iron deficiency at baseline.

**Figure 1.**
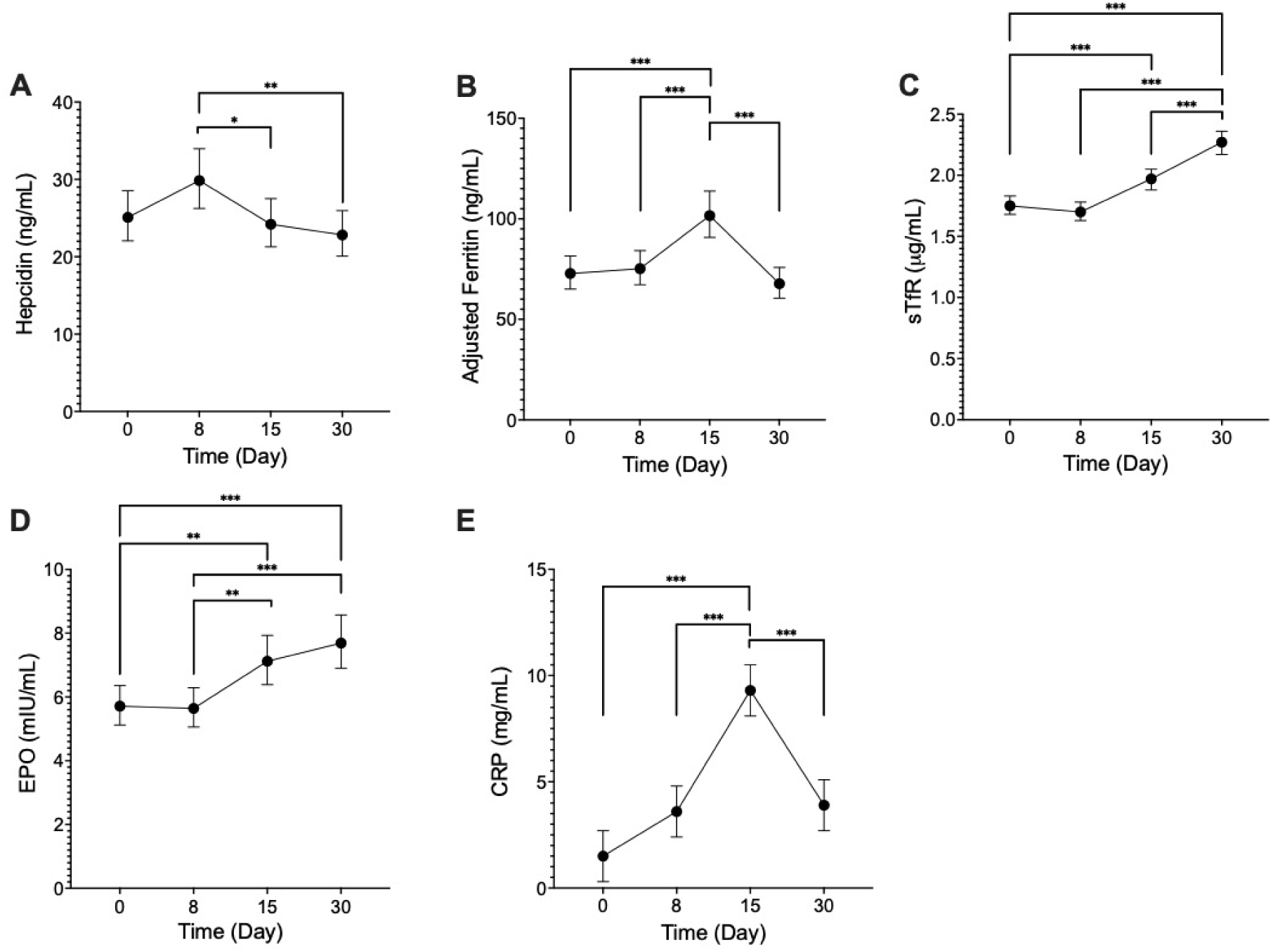
Longitudinal changes in the markers of iron metabolism in 55 volunteers experimentally infected with *P. falciparum*. Data points represent the means, and error bars the 95% confidence intervals. A repeated measures ANOVA was performed on log10 transformed data which were back-transformed to original scale for presentation. Ferritin is adjusted as per the BRINDA project.^27,28^ *p-value <0.05, **p<0.01, ***p<0.001.

Peak hepcidin (day 8) correlated with baseline hepcidin (r=0.76, p<0.001) and baseline adjusted ferritin (r=0.28, p=0.039), although not pre-treatment total parasite burden (**Table 2**). The correlation between baseline and peak hepcidin remained significant on the multivariable model (p<0.001).

Regarding predictors of iron parameters at the end of study (day 30), day 30 sTfR was strongly associated with the baseline sTfR (r=0.85, p<0.0001), and inversely associated with baseline MCV (r=-0.42, p=0.002) and peak hepcidin (r=-0.30, p=0.025). There was no correlation between day 30 sTfR and either day 30 EPO or day 30 reticulocyte count. In the multivariable analysis, the only predictor of day 30 sTfR was baseline sTfR (p<0.001). An increase in reticulocytes was observed from baseline to end of study (median 14 x10^9^/L, [IQR 3-20]; n=39, p<0.001), with this increase associated with baseline ferritin (r=0.34, p=0.033).

### Longitudinal changes in liver transaminases

Median ALT remained unchanged between day 0 (20 U/L [IQR 18-22]) and day 8 (19 U/L [IQR 17-21]) but was transiently increased on day 15 (41 U/L [IQR 37-46]; p<0.001), returning to baseline at EOS (**Figure 2**). Median AST was also transiently increased on day 15 compared to baseline and day 8 levels. The bilirubin did not vary across the duration of the study, with no significant changes.

**Figure 2.**
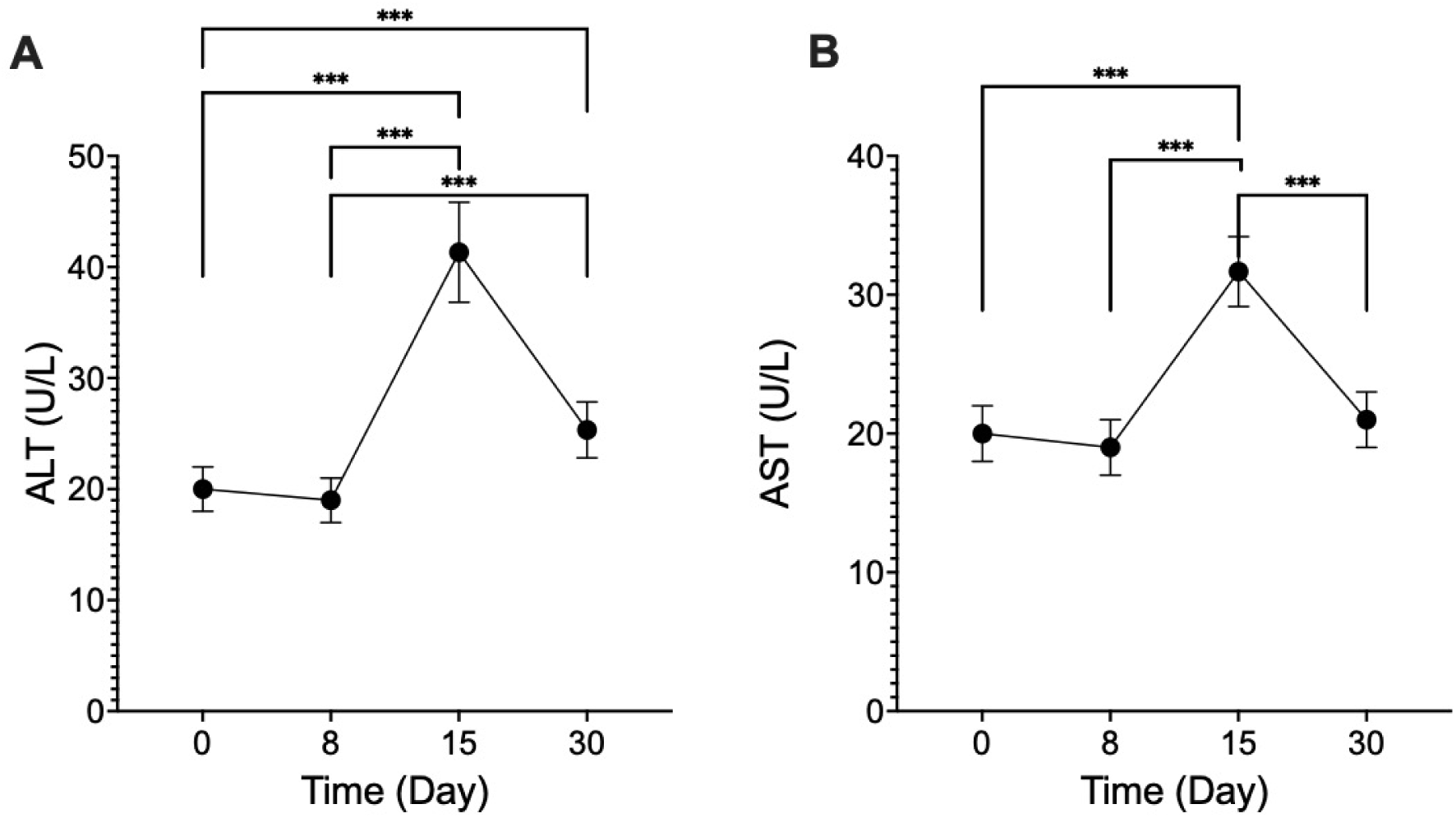
Longitudinal changes in the markers of liver transaminases in 55 volunteers experimentally infected with *P. falciparum*. Data points represent the means, and error bars the 95% confidence intervals. A repeated measures ANOVA was performed on log10 transformed data which were back-transformed to original scale for presentation. *p <0.05, **p<0.01, ***p<0.001.

Day 15 ALT and AST were both associated with baseline adjusted ferritin (ALT: r=0.51, p<0.001; AST: r=0.43, p=0.001) as well as the day 8 adjusted ferritin (ALT: r=0.48, p<0.001; AST: r=0.42, p=0.002) and day 15 adjusted ferritin (ALT: r=0.52, p<0.001; AST: r=0.52, p<0.001) (**Table 3**). Hepcidin and sTfR were not significantly associated with ALT and AST, at any timepoints. Day 15 ALT and AST were also associated with day 15 CRP levels (ALT: r=0.26, p=0.042; AST: r=0.28, p=0.041), as well as with total parasite burden pre-treatment (ALT: r=0.35, p=0.009; AST: r=0.33, p=0.013), although not with peak parasitaemia. In multivariable analysis, total parasite burden pre-treatment (p=0.041) and baseline ferritin (p=0.001) remained significantly correlated with day 15 ALT.

**Table 3.** Associations between markers of iron metabolism and liver enzymes in volunteers experimentally infected with *P. falciparum*. Multivariable linear regression analysis using backward stepwise elimination of log-transformed biological plausible factors. Non-significance for inclusion in the regression model was deemed p>0.20, and variables eliminated until all variables were significant (p<0.05). R^2^: coefficient of determination of the model. ALT: alanine transferase; CRP: C-reactive protein.

| Association with day 15 ALT | Spearman correlation | | Stepwise regression model ($R^2=0.34$) | |
| --- | --- | --- | --- | --- |
|  | R value | P value | Co-efficient (95% confidence interval) | P value |
| Baseline adjusted ferritin | **0.51** | **<0.001** | **0.39 (0.16- 0.62)** | **0.001** |
| Total parasite burden | **0.35** | **0.009** | **0.13 (0.05-0.25)** | **0.041** |
| Day 15 CRP | 0.26 | 0.042 |  |  |
| Peak hepcidin | -0.03 | 0.84 |  |  |

### Malaysian malaria patients

#### Demographics and admission parasitaemia

A total of 201 patients were enrolled and had iron parameters evaluated, including 109 with falciparum and 62 with vivax malaria, as well as 30 healthy controls. Appropriate samples were collected on days 0, 7 and 28 from 33 patients with falciparum and 56 with vivax malaria, with the others having samples collected on days 0 and 28. The median age was 28 years (IQR 19-43, range 7 – 72) for patients with falciparum malaria, and 18 years (IQR 10-35, range 5 - 66) for those with vivax malaria (**Table 4**). Most patients were male (73% [n=79/109] of those with falciparum malaria and 76% [n=47/62] of those with vivax malaria). The median parasitaemia on presentation was higher in patients with *P. falciparum* (6,783 parasites/μL [IQR 1,836-20,020]) compared to those with *P. vivax* (3,629 parasites/μL [IQR 1,226-8,832]). There was no correlation between parasitaemia and MCV on admission in either falciparum or vivax malaria.

**Table 4.** Clinical and laboratory characteristics of Malaysian patients with falciparum and vivax malaria. Variables are median (interquartile range). Pf: *P. falciparum*; Pv: *P. vivax*; sTfR: soluble transferrin receptor.

|  | Control<br>(n=30) | All<br>(n=109) | *P. falciparum* |  | P value<br>(male vs<br>female) | *P. vivax* |  | Female<br>(n=15) | P value<br>(male vs<br>female) | P value<br>(malaria vs<br>control) | P value<br>(Pf vs Pv) |
| --- | --- | --- | --- | --- | --- | --- | --- | --- | --- | --- | --- |
|  |  |  | Male (n=79) | Female<br>(n=30) |  | All<br>(n=62) | Male<br>(n=47) |  |  |  |  |
| Age, years | 36<br>(27-43) | 28<br>(19-43) | 26<br>(19-39) | 38<br>(20-52) | 0.13 | 18<br>(10-35) | 24<br>(11-36) | 11<br>(9-18) | **0.028** | **0.012** | **0.007** |
| Haemoglobin, g/L |  | 131<br>(113-144) | 137<br>(115-146) | 120<br>(104-130) | **0.001** | 124<br>(104-133) | 124<br>(109-139) | 108<br>(93-121) | **0.007** |  | **0.016** |
| Mean<br>corpuscular<br>volume, fL |  | 79<br>(71-86)<br>[n=65] | 79<br>(71-87)<br>[n=47] | 78<br>(68-88)<br>[n=18] | 0.71 | 81<br>(75-88)<br>[n=59] | 84<br>(77-88)<br>[n=45] | 78<br>(71-83)<br>[n=12] | **0.024** |  | 0.23 |
| Parasitaemia on<br>presentation |  | 6,783<br>(1,836-20,020) | 6,848<br>(1,732-21,760) | 6,161<br>(1,917-17,263) | 0.71 | 3,629<br>(1,226-8,832) | 3,739<br>(1,226-8,832) | 3,518<br>(625-9,736) | 0.69 |  | **0.033** |
| Admission<br>hepcidin, ng/mL | 22<br>(16-31) | 136<br>(80-221) | 139<br>(80-216) | 120<br>(69-235) | 0.53 | 130<br>(54-205) | 139<br>(64-224) | 87<br>(30-154) | 0.08 | **<0.001** | 0.21 |
| Day 28 hepcidin,<br>ng/mL |  | 29<br>(13-45) | 33<br>(16-48) | 22<br>(11-39) | 0.05 | 19<br>(12-37) | 22<br>(12-43) | 17<br>(14-27) | 0.38 | 0.46 | 0.13 |
| Admission<br>ferritin, ng/mL | 53<br>(20-90) | 301<br>(202-430) | 330<br>(203-501) | 225<br>(163-334) | **0.016** | 209<br>(146-313) | 207<br>(146-313) | 220<br>(143-351) | 0.64 | **<0.001** | **0.006** |
| Day 28 ferritin,<br>ng/mL |  | 127<br>(52-201) | 152<br>(71-241) | 70<br>(29-112) | **0.001** | 84<br>(35-142) | 87<br>(35-159) | 71<br>(27-95) | 0.32 | **0.003** | **0.023** |
| Admission sTfR,<br>μg/mL | 1.24<br>(1.11-1.57) | 1.24<br>(0.96-1.45) | 1.23<br>(0.95-1.42) | 1.39<br>(1.01-1.70) | 0.10 | 1.24<br>(1.04-1.60) | 1.21<br>(1.00-1.59) | 1.31<br>(1.07-1.70) | 0.46 | 0.44 | 0.44 |
| Day 28 sTfR,<br>μg/mL |  | 1.99<br>(1.63-2.42) | 1.91<br>(1.63-2.27) | 2.18<br>(1.61-2.60) | 0.19 | 2.02<br>(1.57-2.42) | 2.01<br>(1.44-2.36) | 2.13<br>(1.66-2.59) | 0.26 | **<0.001** | 0.99 |

#### Markers of iron metabolism in patients with falciparum and vivax malaria

Hepcidin on admission was significantly higher in malaria patients compared to the healthy controls (median 136 ng/mL [IQR 74-214] vs 22 ng/mL [IQR 16-31]; p<0.001; **Table 4**), and in patients with falciparum malaria was associated with parasitaemia (r=0.36, p=0.001; **Supplementary Table 2**). By day 28, hepcidin in malaria patients had returned to similar levels as the control participants.

The median ferritin on admission was also significantly higher in patients with malaria compared to the controls (245 ng/mL [IQR 172-385] vs 53 ng/ml [IQR 20 – 90]; p<0.001) and was higher in patients with falciparum compared to vivax malaria (301 ng/mL [IQR 202-430] vs 209 ng/mL [146-313]; p=0.006). By day 15, ferritin increased in those treated for vivax but not falciparum malaria. By day 28, median ferritin had fallen significantly in patients with vivax and falciparum malaria (**Figure 3**), but remained more than double that of controls (108 ng/mL [IQR 46-179] vs 53 ng/mL [IQR 20-90]; p=0.003, **Table 4**). There were no associations in either species between day 0 or day 28 ferritin and parasitaemia (**Supplementary Table 2**).

**Figure 3.**
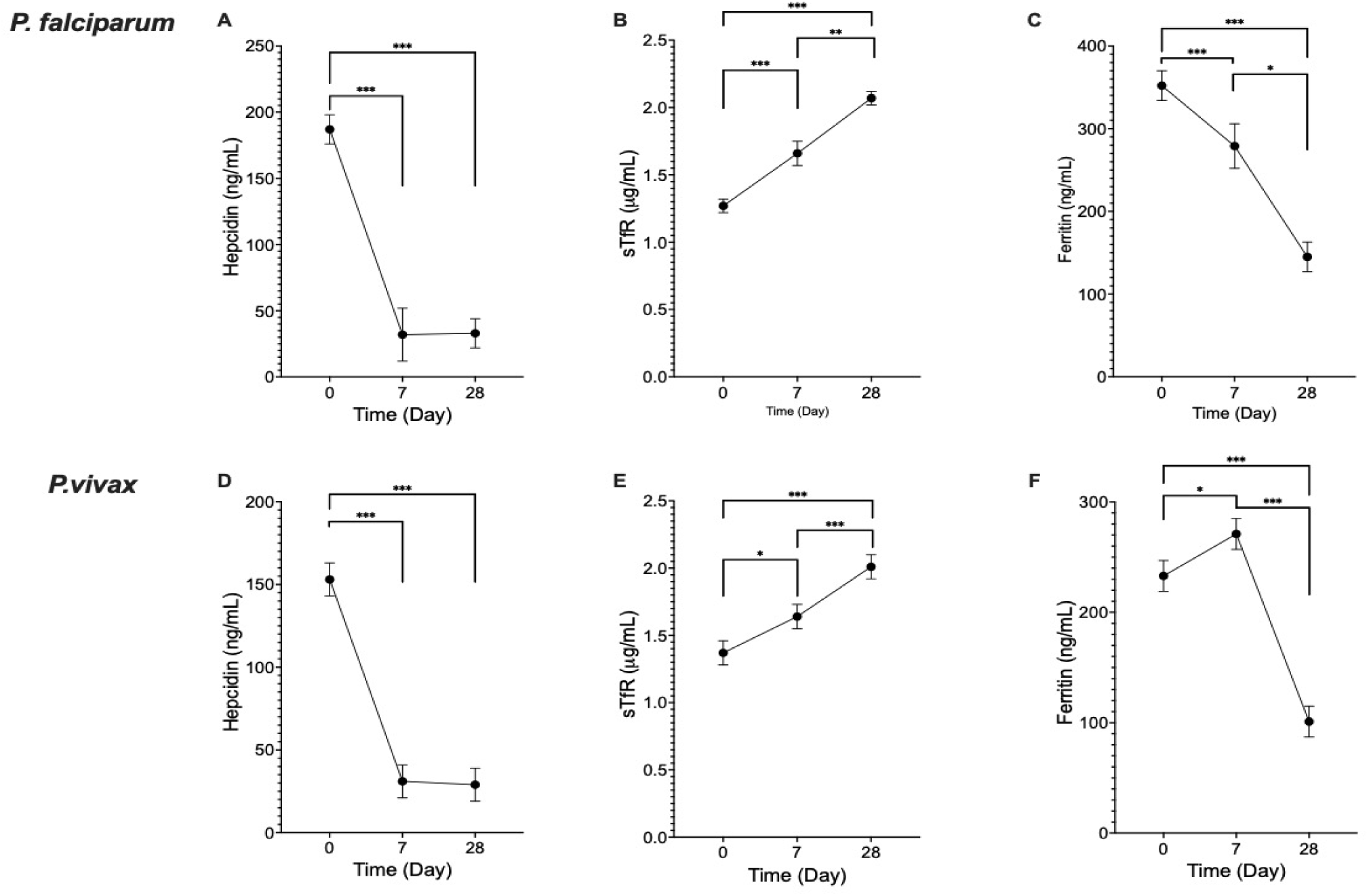
Longitudinal changes in the markers of iron metabolism in Malaysian patients with *P. falciparum* (n=109) *and P. vivax* (n=62) malaria. Data points and error bars represent marginal means and 95% confidence intervals, respectively. Data were analysed using a linear mixed effects model, as day 7 samples were available for only 33/109 patients with falciparum malaria, and 56/62 patients with vivax malaria. Non-normally distributed variables were log10 transformed, and back-transformed for presentation. sTfR: soluble transferrin receptor. *p<0.05, **p<0.01, ***p<0.001.

In patients with malaria, the sTfR levels on admission were similar to those of the controls, however were significantly higher than the controls by day 30 (median 2.00 μg/mL [IQR 1.60-2.42] vs 1.24 μg/mL [IQR 1.11-1.57]; p<0.001). Despite the similar levels of sTfR between malaria patients on admission and controls, sTfR levels were inversely associated with parasitaemia (r=-0.20, p=0.042) in patients with falciparum malaria, possibly reflecting decreased erythropoiesis in patients with higher parasitaemia.

## Discussion

In this study we evaluated the longitudinal changers in markers of iron metabolism in volunteers experimentally infected with blood-stage *P. falciparum*, and in Malaysian patients with falciparum and vivax malaria, to better understand the complex interaction between *Plasmodium* infection and iron homeostasis. We demonstrated that in human experimental malaria parasite growth was not associated with participants’ baseline iron status but was associated with their MCV. We found that hepcidin levels increased in experimental and clinical infection but fell rapidly following treatment, while sTfR remained elevated following recovery in both experimental and clinical malaria. These findings highlight the prolonged disturbance that occurs in iron biomarkers during malaria and extend previous studies in African children^6^ suggesting a possible causal link between malaria infection and functional iron deficiency.

Many pathogens require iron for growth, development, and pathogenicity, and hence it has long been postulated that iron deficiency may be protective against severe manifestations of malaria. Indeed, previous studies have demonstrated that iron deficiency is associated with a reduced risk of malaria in African children^1^ and pregnant women,^34,35^ and that iron supplementation increases risk of malaria infection and mortality.^3^ Furthermore, *in vitro* studies of parasite growth in red blood cells (RBCs) from iron replete donors have demonstrated increased parasite growth,^5^ and a modelling study has suggested an association between host iron bioavailability and parasite multiplication rate.^36^

In contrast to these studies, in the current study where parasite growth rates were evaluated in 55 experimentally infected volunteers, we did not find any correlation between any of the iron parameters at baseline with either peak parasitaemia, total pre-treatment parasite burden, or parasite multiplication rate. We did however find a positive correlation between RBC mean corpuscular volume (MCV) and parasitaemia, with MCV being associated with total pre-treatment parasite burden on multivariable analysis. This finding is consistent with the above-mentioned study by Clark et al. in which parasite growth was increased in RBCs from iron replete donors (all of whom had an MCV >80 fL), compared to RBCs from iron-deficient donors (with an MCV <80 fL).^5^ Furthermore, Clark et al. reported that *P. falciparum* growth and invasion rates increased when old RBCs (which have lower MCV) were replaced with young RBCs (with a higher MCV).^5^ These findings are also consistent with ex-vivo findings in Gambian children with anaemia; multivariable analysis demonstrated that MCV and haemoglobin, but not other markers of iron status, were independently associated with parasite growth.^37^ Taken together, these findings suggest that the protective effect of iron deficiency on clinical malaria and parasitaemia observed in clinical studies may be mediated through the effect of iron deficiency on the structure of the RBCs, rather than by depletion of the iron available to the parasite.

Our study in experimentally infected volunteers provided a unique opportunity to evaluate the longitudinal changes in iron biomarkers that occur in early malaria infection. One previous study has evaluated iron biomarkers in experimental human malaria infection; however this study was small, reporting data from only 5 participants with sporozoite-induced malaria.^38^

In the current study we demonstrate that hepcidin increases very early in infection, peaking at day 8 following inoculation with blood-stage *P. falciparum*, when parasitaemia levels were only just at the threshold for microscopic detection. This is consistent with a previous study by de Mast et al., who demonstrated that hepcidin concentrations were also elevated in Indonesian children with asymptomatic *P. falciparum* and *P. vivax* parasitaemia, despite low CRP levels in these children.^11^ The mechanism of elevated hepcidin at these low parasitaemia levels is uncertain. Hepcidin is known to be produced by the liver in response to IL-6,^10^ and it is possible that the early increase in hepcidin reflects an early inflammatory response. However, we found that CRP was only very mildly elevated by day 8, and like other studies,^39^ we did not find an association between CRP and hepcidin. IL-6 independent mechanisms of hepcidin regulation have been reported, including production of hepcidin by peripheral blood mononuclear cells stimulated by parasitised RBCs,^40^ and may account for the early increase in hepcidin observed in the malaria VIS.

In contrast to the early increase in hepcidin, in experimental malaria infection ferritin did not rise until after treatment, with the rise coinciding with a rapid fall in hepcidin. While ferritin and hepcidin had both returned to baseline by the end of study in experimental malaria, sTfR concentrations rose markedly after treatment, and were at peak levels by the end of study. This may reflect erythropoiesis, with erythropoietin and the reticulocyte count also both increased by the end of study. However, we did not observe any correlation between day 30 sTfR levels and either erythropoietin or reticulocyte count, possibly suggesting that the increased sTfR may also reflect a hepcidin-mediated reduction in total iron body content. Hepcidin depletes available iron by reducing gastrointestinal absorption as well as preventing release of iron from intracellular sources. A hepcidin-mediated iron deficiency resulting from malaria has been suggested by Muriuki et al.,^29^ who evaluated sickle cell trait (a genetic variant protective against malaria) in Mendelian randomization analyses and demonstrated that sickle cell trait was associated with a reduced prevalence of iron deficiency in African children. Our findings from the malaria VIS suggest that hepcidin-mediated iron deficiency following malaria may occur in early primary infections, even at very low parasitaemia levels.

It was notable in our malaria VIS that sTfR concentrations at end of study were strongly correlated with the sTfR concentrations at baseline. While not unexpected, this highlights the fact that individuals who are iron deficient, although perhaps at lower risk of acquiring malaria, may nonetheless be most vulnerable to further depletion in iron stores. We also noted a correlation between baseline ferritin and magnitude of reticulocyte response on recovery, suggesting that individuals with iron deficiency may be at greater risk of malarial anaemia due to impaired erythropoiesis.

In the current study, we also evaluated longitudinal changes of iron parameters in Malaysian patients (mostly adults) with falciparum and vivax malaria. To our knowledge this is the first longitudinal analysis of iron parameters in adults with clinical malaria, although elevated hepcidin has been reported in adults with vivax malaria.^13^ In the current study the temporal changes in the iron biomarkers in Malaysian patients were generally consistent with those of the VIS, and as previously reported in African children. As expected, hepcidin and ferritin were both elevated on admission, and were higher than the levels observed in the VIS. As in the VIS, hepcidin levels fell rapidly following antimalarial treatment. The post-treatment increase in ferritin observed in the VIS was observed in clinical vivax but not falciparum malaria, possibly reflecting a greater post-treatment inflammatory response seen with this *P. vivax.*^41^ In both species ferritin had reduced by day 28, although remained more than twice the level of healthy controls. This is consistent with a previous study by Castberg et al., which demonstrated that in African children ferritin remained elevated up to 4 weeks following malaria infection (when compared to the 6 week post-treatment and presumed baseline level).^15^ Whilst it seems likely that in the current study ferritin may have declined further beyond 28 days, we cannot exclude the possibility that, compared to controls, the malaria patients had increased ferritin prior to infection, and may have been more susceptible to malaria infection, consistent with the above-mentioned studies reporting increased susceptibility to malaria infection in more iron-replete individuals.

As with the VIS, sTfR remained significantly elevated at day 30 in Malaysian patients with clinical malaria, and again raises the possibility that a hepcidin-mediated functional iron deficiency following malaria may also occur in adults. This persistent elevation of both ferritin and sTfR up to one month following malaria infection in adults also suggests that, as with children, these parameters may be unhelpful in determining iron status in adults in malaria endemic regions. In contrast, hepcidin returns rapidly to baseline, and may be a more informative marker.

Finally in this study we evaluated a possible link between iron status and post-treatment elevation in the liver transaminases, alanine transferase (ALT) and aspartate transferase (AST). Post-treatment elevation in ALT/AST are common in malaria VIS^19,20^ and in clinical malaria in non-immune patients.^21^ In malaria VIS these transient elevations in liver transaminases have the potential to be misinterpreted as being attributable to the investigational antimalarial, thereby inappropriately halt drug development. Thus, understanding the factors contributing to these post-treatment changes are important for antimalarial drug development. Post-treatment transaminitis in malaria VIS has previously been attributed to inflammation, with peak ALT associated with peak CRP.^20^ However, in this study we also found that baseline ferritin strongly predicted day 15 ALT/AST, even after controlling for other contributing factors such as parasitaemia and CRP. We hypothesise that this post-treatment increase in ALT/AST may reflect a hepatic immune response, with this response more exaggerated in those who are more iron replete and thus have a more robust immune response. This latter hypothesis is consistent with a recent study demonstrating attenuated hepatic immune response and reduced hepatic pathology in *P. chabaudi*-infected transgenic mice carrying a mutation in the transferrin receptor causing decreased cellular iron uptake.^42^

Our study had several limitations. The sample sizes in both the malaria VIS and the Malaysian studies were relatively small, and it is possible that we were underpowered to detect significant correlations. In both the malaria VIS and the Malaysian studies follow up was limited to approximately 4 weeks, and thus we were unable to determine any changes beyond this time. Finally, CRP was not measured in the Malaysian patients, and so we were unable to adjust ferritin for CRP.

In summary, our studies provide a detailed description of the longitudinal changes that occur in iron homeostasis during a malaria infection. We demonstrate that in experimental infection MCV, rather than iron status, is associated with parasitaemia, and demonstrate the very early increase in hepcidin in blood stage malaria infection. Importantly we provide information on the duration of the disturbance in iron homeostasis following recovery from clinical malaria in adults, demonstrating that while hepcidin declines rapidly to the level of healthy controls, ferritin and sTfR both remain above the levels of healthy controls up to 1 month post recovery. Finally, we demonstrate an association between iron status and risk of post-treatment elevations in liver transaminases in malaria VIS, providing insights into the potential mechanisms of this finding, and providing further reassurance that these changes are a common response to treatment of malaria rather than specific drug effect.

## Data Availability

Data will be made available upon request through contact with the corresponding author, with an appropriate data sharing agreement in place.

## Contributors

S.D.W., J.S.M., N.M.A. and B.E.B. conceived and designed the study. B.E.B, M.J.G, T.W. and G.S.R. conducted the clinical studies Malaysia. S.D.W., B.E.B., and J.S.M, conducted the malaria volunteer infection studies. K.P., J.G., A.S.N., and F.M.A. conducted the assays. S.D.W., L.M. and B.E.B. analysed the data. S.D.W. and B.E.B. wrote the paper. All authors reviewed the manuscript, and all approved the final draft of the manuscript. S.D.W. and B.E.B have accessed and verified the data.

## Acknowledgements

We thank all volunteers enrolled in the malaria volunteer infection studies in Brisbane, Australia, and the patients enrolled in the clinical studies in Malaysia; research and clinical staff at the participating hospitals in Malaysia; and clinical staff at QPharm and University of Sunshine Coast Clinical Trial Sites, Brisbane, Australia.

This study was supported by the National Health and Medical Research Council (Program Grant 1037304, Project Grants 1045156 and 1156809; Investigator Grants 2016792 to BEB, 2016396 to JCM, 2017436 to MJG), the US National Institute of Health (R01 AI116472-03), and the Malaysian Ministry of Health (Grant BP00500420). The malaria volunteer infection studies were funded by Medicines for Malaria Venture, Cadila Healthcare Ltd, the Bill and Melinda Gates Foundation (INV-001965), and the Global Health Innovative Technology Fund (Grant G2016-226).

## References

1. Muriuki JM, Mentzer AJ, Kimita W, et al. Iron status and associated malaria risk among African children. Clin Infect Dis 2019; 68: 1807–14.

2. Kabyemela ER, Fried M, Kurtis JD, Mutabingwa TK, Duffy PE. Decreased susceptibility to *Plasmodium falciparum* infection in pregnant women with iron deficiency. J Infect Dis 2008; 198: 163–6.

3. Gwamaka M, Kurtis JD, Sorensen BE, et al. Iron deficiency protects against severe *Plasmodium falciparum* malaria and death in young children. Clin Infect Dis 2012; 54: 1137–44.

4. Sazawal S, Black RE, Ramsan M, et al. Effects of routine prophylactic supplementation with iron and folic acid on admission to hospital and mortality in preschool children in a high malaria transmission setting: community-based, randomised, placebo-controlled trial. Lancet 2006; 367: 133–43.

5. Clark MA, Goheen MM, Fulford A, et al. Host iron status and iron supplementation mediate susceptibility to erythrocytic stage *Plasmodium falciparum*. Nat Commun 2014; 5: 4446.

6. Muriuki JM, Mentzer AJ, Mitchell R, et al. Malaria is a cause of iron deficiency in African children. Nat Med 2021; 27: 653–8.

7. Drakesmith H, Prentice AM. Hepcidin and the iron-infection axis. Science 2012; 338: 768–72.

8. Anderson GJ, Frazer DM. Current understanding of iron homeostasis. Am J Clin Nutr 2017; 106: 1559S–66S.

9. Knutson MD. Iron transport proteins: Gateways of cellular and systemic iron homeostasis. J Biol Chem 2017; 292: 12735–43.

10. Pietrangelo A, Dierssen U, Valli L, et al. STAT3 is required for IL-6-gp130–dependent activation of hepcidin in vivo. Gastroenterol 2007; 132: 294–300.

11. de Mast Q, Syafruddin D, Keijmel S, et al. Increased serum hepcidin and alterations in blood iron parameters associated with asymptomatic *P. falciparum* and *P. vivax* malaria. Haematologica 2010; 95: 1068.

12. Kaboré B, Post A, Berendsen ML, et al. Red blood cell homeostasis in children and adults with and without asymptomatic malaria infection in Burkina Faso. PLoS ONE 2020; 15: e0242507.

13. Mendonça VR, Souza LC, Garcia GC, et al. Associations between hepcidin and immune response in individuals with hyperbilirubinaemia and severe malaria due to *Plasmodium vivax* infection. Malar J 2015; 14: 1–11.

14. Barffour MA, Schulze KJ, Coles CL, et al. Malaria exacerbates inflammation-associated elevation in ferritin and soluble transferrin receptor with only modest effects on iron deficiency and iron deficiency anaemia among rural Zambian children. Trop Med Int Health 2018; 23: 53–62.

15. Castberg FC, Sarbah EW, Koram KA, et al. Malaria causes long-term effects on markers of iron status in children: a critical assessment of existing clinical and epidemiological tools. Malar J 2018; 17: 1–12.

16. Suominen P, Punnonen K, Rajamäki A, Irjala K. Serum transferrin receptor and transferrin receptor-ferritin index identify healthy subjects with subclinical iron deficits. Blood 1998; 92: 2934–9.

17. Ntenda PA, Chirambo AC, Nkoka O, El-Meidany WM, Goupeyou-Youmsi J. Implication of asymptomatic and clinical *Plasmodium falciparum* infections on biomarkers of iron status among school-aged children in Malawi. Malar J 2022; 21: 278.

18. Menendez C, Quinto L, Kahigwa E, et al. Effect of malaria on soluble transferrin receptor levels in Tanzanian infants. Am J Trop Med Hyg 2001; 65: 138–42.

19. Chughlay MF, Akakpo S, Odedra A, et al. Liver enzyme elevations in *Plasmodium falciparum* volunteer infection studies: findings and recommendations. Am J Trop Med Hyg 2020; 103: 378.

20. Odedra A, Webb L, Marquart L, et al. Liver function test abnormalities in experimental and clinical *Plasmodium vivax* infection. Am J Trop Med Hyg 2020.

21. Woodford J, Shanks GD, Griffin P, Chalon S, McCarthy JS. The dynamics of liver function test abnormalities after malaria infection: a retrospective observational study. Am J Trop Med Hyg 2018; 98: 1113.

22. Gaur AH, McCarthy JS, Panetta JC, et al. Safety, tolerability, pharmacokinetics, and antimalarial efficacy of a novel *Plasmodium falciparum* ATP4 inhibitor SJ733: a first-in-human and induced blood-stage malaria phase 1a/b trial. Lancet Infect Dis 2020; 20: 964–75.

23. Barber BE, Fernandez M, Patel H, et al. Safety, pharmacokinetics and antimalarial activity of the novel triaminopyrimidine ZY-19489: a first-in-human, randomised, placebo-controlled, double-blind, single ascending dose study, a pilot food effect study, and a volunteer infection study. Lancet Infect Dis 2022; 22: 879–90.

24. Woolley SD, Fernandez M, Rebelo M, et al. Development and evaluation of a new *Plasmodium falciparum* 3D7 blood stage malaria cell bank for use in malaria volunteer infection studies. Malar J 2021; 20: 1–9.

25. Barber BE, Abd-Rahman A, Webster R, et al. Characterizing the blood stage antimalarial activity of tafenoquine in healthy volunteers experimentally infected with *Plasmodium falciparum*. Clin Infect Dis 2023; 76: 1919–27.

26. Watts R, Odedra A, Marquart L, et al. Safety and parasite clearance of artemisinin-resistant *Plasmodium falciparum* infection: A pilot and a randomised volunteer infection study in Australia. PLoS Med 2020; 17: e1003203.

27. Suchdev PS, Williams AM, Mei Z, et al. Assessment of iron status in settings of inflammation: challenges and potential approaches. Am J Clin Nutr 2017; 106: 1626S–33S.

28. Namaste SM, Rohner F, Huang J, et al. Adjusting ferritin concentrations for inflammation: Biomarkers Reflecting Inflammation and Nutritional Determinants of Anemia (BRINDA) project. Am J Clin Nutr 2017; 106: 359S–71S.

29. Barber BE, William T, Grigg MJ, et al. A prospective comparative study of knowlesi, falciparum and vivax malaria in Sabah, Malaysia: high proportion with severe disease from *Plasmodium knowlesi* and *P. vivax* but no mortality with early referral and artesunate therapy. Clin Infect Dis 2013; 56: 383–97.

30. Barber BE, Grigg MJ, Piera K, et al. Effects of aging on parasite biomass, inflammation, endothelial activation and microvascular dysfunction in *Plasmodium knowlesi* and *P. falciparum* malaria J Infect Dis 2017; 215: 1908–17.

31. Grigg MJ, William T, Barber BE, et al. Artemether-lumefantrine versus chloroquine for the treatment of uncomplicated *Plasmodium knowlesi* malaria: an open-label randomized controlled trial CAN KNOW. Clin Infect Dis 2018; 66: 229–36.

32. Grigg MJ, William T, Menon J, et al. Artesunate–mefloquine versus chloroquine for treatment of uncomplicated *Plasmodium knowlesi* malaria in Malaysia (ACT KNOW): an open-label, randomised controlled trial. Lancet Infect Dis 2016; 16: 180–8.

33. Grigg MJ, William T, Menon J, et al. Efficacy of artesunate-mefloquine against high-grade chloroquine-resistant *Plasmodium vivax* malaria in Malaysia: an open-label randomised controlled trial. Clin Infect Dis 2016; 62: 1403–11.

34. Senga EL, Harper G, Koshy G, Kazembe PN, Brabin BJ. Reduced risk for placental malaria in iron deficient women. Malar J 2011; 10: 1–5.

35. Unger HW, Bleicher A, Ome-Kaius M, Aitken EH, Rogerson SJ. Associations of maternal iron deficiency with malaria infection in a cohort of pregnant Papua New Guinean women. Malar J 2022; 21: 1–11.

36. Georgiadou A, Lee HJ, Walther M, et al. Modelling pathogen load dynamics to elucidate mechanistic determinants of host–*Plasmodium falciparum* interactions. Nat Microbiol 2019; 4: 1592–602.

37. Goheen M, Wegmüller R, Bah A, et al. Anemia offers stronger protection than sickle cell trait against the erythrocytic stage of falciparum malaria and this protection is reversed by iron supplementation. EBioMed 2016; 14: 123–30.

38. De Mast Q, Van Dongen-Lases EC, Swinkels DW, et al. Mild increases in serum hepcidin and interleukin-6 concentrations impair iron incorporation in haemoglobin during an experimental human malaria infection. Br J Haematol 2009; 145: 657–64.

39. de Mast Q, Nadjm B, Reyburn H, et al. Assessment of urinary concentrations of hepcidin provides novel insight into disturbances in iron homeostasis during malarial infection. J Infect Dis 2009; 199: 253–62.

40. Armitage AE, Pinches R, Eddowes LA, Newbold CI, Drakesmith H. *Plasmodium falciparum* infected erythrocytes induce hepcidin (HAMP) mRNA synthesis by peripheral blood mononuclear cells. Br J Haematol 2009; 147: 769–71.

41. Anstey NM, Douglas NM, Poespoprodjo JR, Price R. *Plasmodium vivax*: clinical spectrum, risk factors and pathogenesis. Adv Parasitol 2012; 80: 151–201.

42. Wideman SK, Frost JN, Richter FC, et al. Cellular iron governs the host response to malaria. PLoS Pathog 2023; 19: e1011679.

