## Supplementary Data for "Longitudinal changes in iron homeostasis in human experimental and clinical malaria"

**Supplementary Table 1. Spearman's correlations between markers of iron metabolism and parasite parameters in the malaria volunteer infection studies.** PMR: parasite multiplication rate; $PP_{Pre}$: peak pre-treatment parasitaemia; sTfR: soluble transferrin receptor; $TPB_{Pre}$: total pre-treatment parasite burden.

| Association | R-value | P-value |
|---|---|---|
| Baseline sTfR with $TPB_{Pre}$ | -0.25 | 0.07 |
| Baseline sTfR with $PP_{Pre}$ | -0.14 | 0.30 |
| Baseline sTfR with PMR | 0.13 | 0.34 |
| Baseline adjusted ferritin with $TPB_{Pre}$ | 0.19 | 0.17 |
| Baseline adjusted ferritin with $PP_{Pre}$ | 0.10 | 0.49 |
| Baseline adjusted ferritin with PMR | 0.10 | 0.51 |

**Supplementary Table 2. Spearman's correlations between markers of iron metabolism and clinical parameters in Malaysian patients.** MCV: mean corpuscular volume; sTfR: soluble transferrin receptor.

| | ***P. falciparum*** **(n=109)** | | ***P. vivax*** **(n=62)** | |
|---|---|---|---|---|
| **Association** | **R score** | **P value** | **R score** | **P value** |
| MCV with parasitaemia | -0.12 | 0.46 | -0.07 | 0.60 |
| Admission sTfR with parasitaemia | **-0.20** | **0.042** | -0.11 | 0.42 |
| Admission hepcidin with parasitaemia | **0.36** | **0.001** | 0.17 | 0.18 |
| Admission ferritin with parasitaemia | 0.07 | 0.49 | 0.06 | 0.65 |
| 28-day ferritin with parasitaemia | 0.07 | 0.50 | 0.03 | 0.65 |
